# Comparison of three bioinformatics tools in the detection of ASD candidate variants from whole exome sequencing data

**DOI:** 10.1101/2023.04.23.23288995

**Authors:** Apurba Shil, Liron Levin, Hava Golan, Gal Meiri, Analya Michaelovski, Yair Tsadaka, Adi Aran, Ilan Dinstein, Idan Menashe

**Affiliations:** Department of Epidemiology, Biostatistics, and Health Community Sciences, Faculty of Health Sciences, Ben-Gurion University of the Negev, Beer-Sheva, Israel; Azrieli National Centre for Autism and Neurodevelopment Research, Ben-Gurion University of the Negev, Beer-Sheva, Israel; The School of Brain Sciences and Cognition, Ben-Gurion University of the Negev, Beer-Sheva, Israel; Bioinformatics Core Facility, Ben-Gurion University of the Negev, Beer-Sheva, Israel; Department of Physiology and Cell Biology, Faculty of Health Sciences, Ben-Gurion University of the Negev, Beer-Sheva, Israel; Preschool Psychiatric Unit, Soroka University Medical Center, Beer-Sheva, Israel; Child Development Center, Soroka University Medical Center, Beer-Sheva, Israel; Child Development Center, Ministry of Health, Beer-Sheva, Israel; Psychology Neuropediatric Unit, Shaare Zedek Medical Center, Jerusalem, Israel; Psychology Department, Ben-Gurion University of the Negev, Beer-Sheva, Israel

**Keywords:** Whole-exome sequencing, Autism Spectrum disorder, Bioinformatics, Genetics, Genomics

## Abstract

**Background:** Autism spectrum disorder (ASD) is a heterogenous multifactorial neurodevelopmental condition with a significant genetic susceptibility component. Thus, identifying genetic variations associated with ASD is a complex task. Whole-exome sequencing (WES) is an effective approach for detecting extremely rare protein-coding single-nucleotide variants (SNVs) and short insertions/deletions (INDELs). However, interpreting these variants’ functional and clinical consequences requires integrating multifaceted genomic information.

**Methods:** We compared the concordance and effectiveness of three bioinformatics tools in detecting ASD candidate variants (SNVs and short INDELs) from WES data of 220 ASD family trios registered in the National Autism Database of Israel. We studied only rare (<1% population frequency) proband-specific variants. According to the American College of Medical Genetics (ACMG) guidelines, the pathogenicity of variants was evaluated by the *InterVar* and *TAPES* tools. In addition, likely gene-disrupting (LGD) variants were detected based on an in-house bioinformatics tool, *Psi-Variant*, that integrates results from seven in-silico prediction tools.

**Results:** Overall, 605 variants in 499 genes distributed in 193 probands were detected by these tools. The overlap between the tools was 64.1%, 17.0%, and 21.6% for *InterVar–TAPES*, *InterVar*– *Psi-Variant*, and *TAPES*–*Psi-Variant*, respectively. The intersection between *InterVar* and *Psi-Variant* (*<u>I</u>*∩*<u>P</u>*) was the most effective approach in detecting variants in known ASD genes (OR = 5.38, 95% C.I. = 3.25–8.53), while the union of *InterVar* and *Psi-Variant* (*<u>I</u>* U *<u>P</u>*) achieved the highest diagnostic yield (30.9%).

**Conclusions:** Our results suggest that integrating different variant interpretation approaches in detecting ASD candidate variants from WES data is superior to each approach alone. The inclusion of additional criteria could further improve the detection of ASD candidate variants.

## Background

Autism spectrum disorder (ASD) comprises a collection of heterogeneous neurodevelopmental disorders that share two behavioral characteristics—difficulties in social communication and restricted, repetitive behaviors and interests^1,2^. The etiology of ASD has a significant genetic component, as is evident from multiple twin and family studies^3–6^. Yet, over the years, very few genetic causes of ASD have been discovered; thus, today, despite extensive research, an understanding of the overall genetic architecture of ASD remains obscure^6,7^.

The emergence of next-generation sequencing (NGS) approaches in the past decade has transformed the genetic research of complex traits^8^. These NGS technologies have facilitated high-throughput DNA sequencing for large cohorts of patients, allowing the comparison of multiple variants that includes single-nucleotide variants (SNVs) and short insertions/deletions (INDELs) between large groups of patients^9–12^. In this realm, whole-exome sequencing (WES) is particularly suitable for studying the genetics of heterogenous traits such as ASD, as it focuses on a relatively limited number of protein-coding variants^9,10,13–18^.

However, understanding the functional consequences of coding Variants is not a trivial task. In 2015, the American College of Medical Genetics and Genomics (ACMG) and the Association for Molecular Pathology (AMP) published standards and guidelines to generalize sequence variant interpretation and to address the issue of inconsistent interpretation across laboratories^8^. The resulting system for classifying variants recommends 28 criteria (16 for pathogenic and 12 for benign variants) and provides a set of scoring rules based on variant population allele frequency, variant functional annotation, variant familial segregation, etc.^8,19^; Variants are classified as pathogenic (P), likely pathogenic (LP), variants of uncertain significance (VUS), likely benign (LB) or benign (B). Subsequently, multiple in-silico tools were developed to implement these ACMG/AMP criteria for annotating the prospective pathogenicity of variants detected in WES studies.

While the ACMG/AMP scoring approach is highly effective for detecting de-novo highly penetrant mutations for rare Mendelian disorders, it is less suitable for detecting inherited partially penetrant variants^20^. Such variants, usually annotated as VUS in terms of the ACMP/AMP criteria, are expected to contribute significantly to the risk of developing neurodevelopmental conditions, including ASD^9,17,18,21,22^. Thus, relying solely on the ACMG/AMP criteria for variant annotation in WES studies of ASD may result in an under-representation of susceptibility variants, which will lead to a lower diagnostic yield for ASD. To overcome this potential limitation, we have developed *“Psi-Variant*,” a pipeline to detect different types of likely gene-disrupting (LGD) variants, including protein truncating and deleterious missense variants. We applied *Psi-Variant* – in comparison with *InterVar* and *TAPES*, two variant interpretation tools that use the ACMG/AMP criteria – to a large WES dataset of ASD to evaluate the concordance between these tools to detect variants and to assess their effectiveness in detecting ASD susceptibility variants.

## Methods

### Study sample

Initially, the study sample comprised 250 children diagnosed with ASD who are registered in the National Autism Database of Israel (NADI)^23,24^ and whose parents gave consent for participation in this study. Based on our clinical records, none of the parents in the study has been diagnosed with ASD, intellectual disability, or any other type of neurodevelopmental disorder. Genomic DNA was extracted from saliva samples from participating children and their parents using Oragene®•DNA (OG-500/575) collection kits (DNA Genotek, Canada).

### Whole exome sequencing

WES analysis was performed on the above-mentioned samples with Illumina HiSeq sequencers, followed by the Illumina Nextera exome capture kit at the Broad Institute as part of the Autism Sequencing Consortium, described previously^11^. Sequencing reads aligned to Genome Reference Consortium Human Build 38 and aggregated into BAM/CRAM files were analyzed using the Genome Analysis Toolkit (GATK)^25^ to generate a joint variant calling format (vcf) file for all subjects in the study. We excluded data for 30 probands from the raw vcf file due to incomplete pedigree information or low-quality WES data. Thus, WES data for 220 ASD trios were analyzed in this study (Fig. 1).

**Fig. 1.**
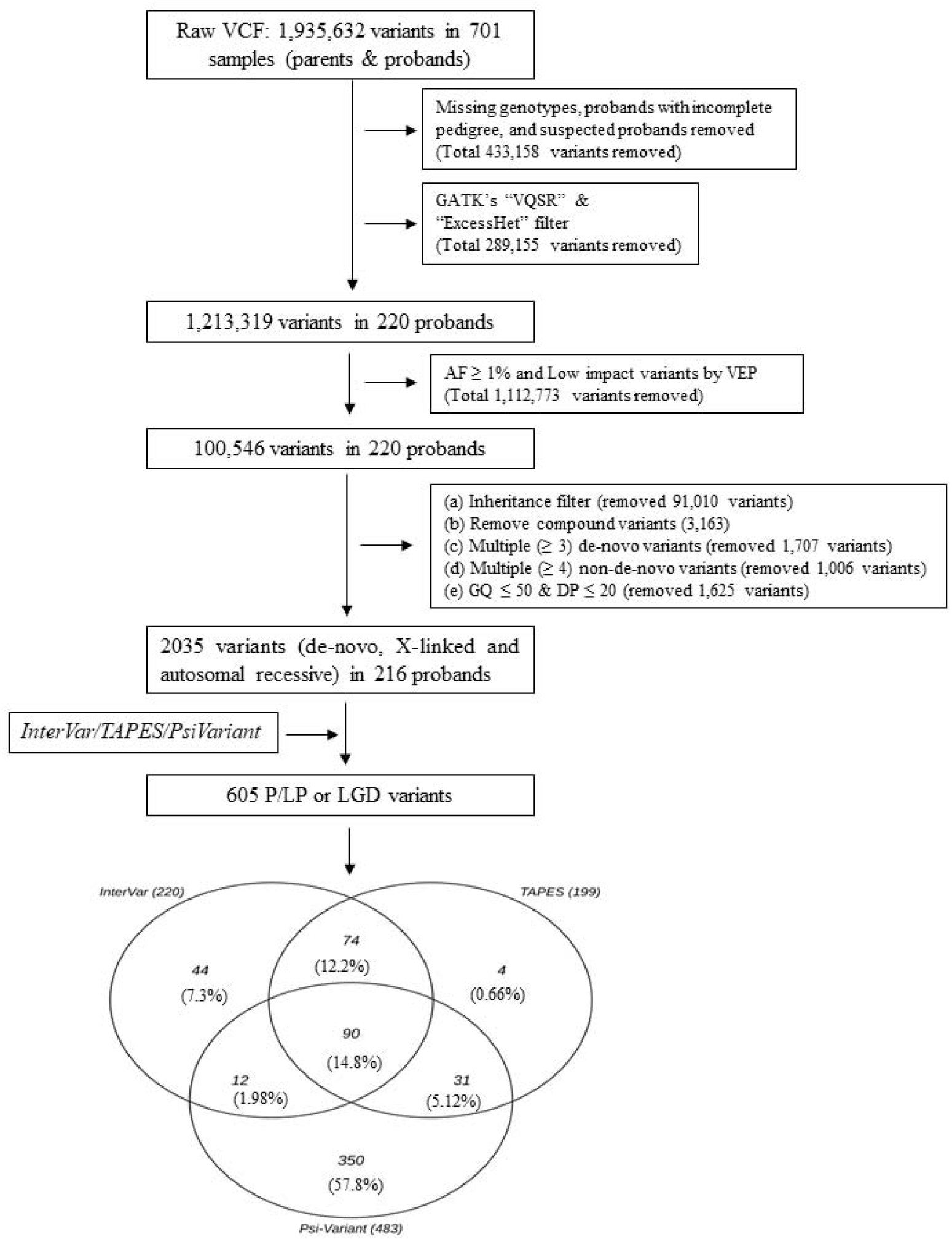
Analysis workflow for detecting LP/P/LGD Variants from the WES data. InterVar and TAPES detected LP/P Variants by implementing ACMG/AMP criteria. *Psi-Variant* detected LGD Variants by utilizing in-house criteria.

### Data analysis

The SNV detection process in this study is outlined in Fig. 1. and explained below.

#### Data cleaning

The raw vcf file contained 1,935,632 variants. From this file, we removed variants with missing genotypes and/or variants in regions with low read coverage (≤ 20 reads) and/or with low genotype quality (GQ ≤ 50). In addition, we removed all common variants (i.e., those with a population minor allele frequency >1%)^26^ as well as those that did not pass the GATK’s “VQSR” and “ExcessHet” filters. Thereafter, we used an in-house machine learning (ML) algorithm to remove other potentially false-positive variants. The details of this ML algorithm and its efficiency in classifying true positive and false positive variants are summarized in the supplementary file S1. Finally, we used the pedigree structure of the families to identify proband-specific genotypes, including de-novo variants, recessively inherited variants, and X-linked variants (in males). Recessively inherited variants occur in the same loci of both copies of a gene in autosomes (where both the parents are carriers). Whereas one altered copy of the gene in chromosome X among males is defined as X-linked (males). We removed multiallelic variants from these genotypes and those classified as “de-novo” that appeared in more than two individuals in the sample. In this study, we haven’t considered compound heterozygote variants (in cis/trans).

#### Identifying ASD candidate variants

We searched for candidate ASD Variants using three complementary approaches. First, we applied *InterVar*^19^ and *TAPES*^27^, two commonly used publicly available command-line tools that use ACMG/AMP criteria^8^, to detect LP/P Variants. In addition, we assigned the ACMG/AMP PS2 criterion to all the de-novo Variants to detect additional LP/P Variants from the list of VUS. Since *InterVar* and *TAPES* are less sensitive tools for detecting recessive possible gene disrupting (LGD) variants^20^, we developed an integrated in-house tool, *Psi-Variant*, to detect LGD variants. The *Psi-Variant* workflow starts using Ensembl’s Variant Effect Predictor (VEP)^26^ to annotate the functional consequences for each variant in a multi-sample vcf file. Then, all frameshift indels, nonsense, and splice acceptor/donor variants are further analyzed by the LoFtool^28^ with scores of < 0.25 are annotated as intolerant variants. In addition, it applies six different in-silico tools to all missense substitutions and annotates them as “deleterious/damaging” if at least three (≥ 50%) of them exceed the following cutoffs: SIFT^29^ (< 0.05), PolyPhen-2^30^ (≥ 0.15), CADD^31^ (> 20), REVEL^32^ (> 0.50), M_CAP^33^ (> 0.025) and MPC^34^ (≥ 2). These scores were extracted by utilizing the dbNSFP database^35^.

#### Comparison between InterVar, TAPES, and Psi-Variant

We compared the number of variants detected by the three tools and the percentages of variants detected by different combinations. Thereafter, we used the list of ASD genes (n = 1031) from the SFARI Gene database^36^ (accessed on 11 January 2022) as the gold standard to compute the odds ratio (OR) and positive predictive value (PPV) for detecting candidate ASD variants in SFARI genes. In addition, we assessed the detection yield for each tool combination by computing the proportion of children with detected candidate ASD variants in SFARI genes.

#### Software

Data storage, management, and analysis were conducted on a high-performing computer cluster in a Linux environment using Python version 3.5 and R Studio version 1.1.456. All the statistical analyses and data visualizations were incorporated into R Studio.

## Results

### Detection of candidate variants by the different tools

A total of 605 variants in 193 probands (highlighted in the supplementary Table S2) were detected by at least one of *InterVar* (n = 220), *TAPES* (n = 199), or *Psi-Variant* (n = 483) from a dataset of 2,035 high-quality, ultra-rare variants with proband-specific genotypes (Fig. 1). Of these, 90 variants (14.9%) were detected by all three tools. The highest concordance in detected variants was observed between *InterVar* and *TAPES* (64.3%), followed by *TAPES* and *Psi-Variant* (21.6%) and *InterVar* and *Psi-Variant* (17.0%).

The characteristics of the detected variants are shown in Table 1. Significantly higher rates of LoF and missense variants were detected by all three tools compared to the rates of these variants in the clean vcf file (*P* < 0.001). As expected, missense variants comprised the majority of detected variants, with 81.6%, 58.8%, and 51.4% of the variants detected by *Psi-Variant, TAPES,* and *InterVar*, respectively. Notably, a higher number of frameshift variants were detected by *Psi-Variant* than by *InterVar* and *TAPES* (58 vs. 39 and 22, respectively), but the percentages of these variants out of the total number of detected variants were lower due to the markedly higher number of variants detected by *Psi-Variant*.

**Table 1.** Characteristics of the detected Variants by *InterVar*, *TAPES*, and *Psi-Variant* from the WES data.

| Characteristics | Preliminary output<br>(n = 1213319) | *InterVar*<br>(n = 220) | *TAPES*<br>(n = 199) | *Psi-Variant*<br>(n = 483) |
| --- | --- | --- | --- | --- |
| **Functional consequence** |  |  |  |  |
| Frameshift (insertions/deletions) | 4232 (0.349%) | 39 (17.7%) * | 22 (11.1%) * | 58 (12.01%) * |
| Missense | 95919 (7.91%) | 113 (51.4%) * | 117 (58.8%) * | 394 (81.6%) * |
| Stop Gain/Loss/retain,<br>Start Gain/Loss | 2105 (0.17%) | 16 (7.27%) * | 13 (6.53%) * | 16 (3.31%) * |
| Non-frameshift/in-frame | 4062 (0.33%) | 42 (19.1%) * | 43 (21.61%) * | -- |
| Splice acceptor/donor/region | 18817 (1.55%) | 4 (1.82%) | 4 (2.01%) | 12 (2.48%) |
| Synonymous, downstream/upstream<br>gene, intron variant | 871205 (71.8%) | 6 (2.73%) | 0 (0%) | 3 (0.62%) |
| Other | 216979 (17.9%) | -- | -- | -- |
| **Inheritance pattern wise** |  |  |  |  |
| De-novo | 43052 (3.55%) | 209 (95%) * | 193 (97%) * | 175 (36.2%) * |
| Autosomal recessive | 70948 (5.85%) | 9 (4.09%) | 5 (2.51%) | 179 (37.1%) * |
| X-linked | 9103 (0.75%) | 2 (0.91%) * | 1 (0.5%) * | 129 (26.7%) * |
| Other | 1090216 (89.8%) | -- | -- | -- |
| **Gene type wise** |  |  |  |  |
| SFARI genes with a score 1 | 19236 (1.58%) | 15 (6.82%) * | 12 (6.03%) * | 21 (4.35%) * |
| All SFARI genes (with scores 1-3) | 93681 (7.72%) | 32 (14.5%) * | 24 (12.1%) * | 75 (15.5%) * |
| Other genes | 1119638 (92.28%) | 188 (85.4%) * | 175 (87.9%) * | 408 (84.5%) * |

\* <0.05 level of significance; two-sided two proportions Z test

Almost all (≥ 95%) variants detected by either *InterVar* or *TAPES* were de-novo variants, while de-novo variants comprised only 36.2% of the variants detected by *Psi-Variant*, which also detected a high portion of X-linked and autosomal recessive variants (37.1% and 26.7%, respectively). Examination of the distribution of the detected variants in genes associated with ASD according to the SFARI Gene database^36^ revealed a two-fold enrichment of variants distributed in ASD genes (for all detection tools) compared to their portion in the clean vcf file and even a higher enrichment of Variants distributed in high-confidence ASD genes (*P* < 0.001).

### Effectiveness of ASD candidate Variants detection

To assess the effectiveness of the different tools in detecting ASD candidate SNVs, we calculated the PPV and the OR for detecting ASD genes (i.e., those listed in the SFARI Gene database^36^) for different combinations of utilization of the three tools. The results of these analyses are shown in Fig. 2. Utilization of any of the three tools resulted in a significant enrichment of ASD genes, with the highest enrichment being observed in SNVs detected by *InterVar* (PPV = 0.178; OR = 4.10, 95% confidence interval (C.I.) = 2.77–5.90) followed by *TAPES* (PPV = 0.158; OR = 3.53, 95% C.I. = 2.28–5.27) and *Psi-Variant* (PPV = 0.143; OR = 3.21, 95% C.I. = 2.39–4.22). Notably, better performance in detecting ASD candidate SNVs was obtained at the intersection of the detected SNVs between *InterVar* and *Psi-Variant* (*<u>I</u>* ∩ *<u>P</u>*) (PPV = 0.222; OR = 5.38, 95% CI = 3.25–8.53). The *<u>I</u>* ∩ *<u>P</u>* combination was also the most effective in detecting SNVs in high-confidence ASD genes (i.e., those with a score of 1 in the SFARI Gene database ^36^ (Fig. 2A -2B). However, the *<u>I</u>* ∩ *<u>P</u>* combination had a relatively low diagnostic yield of 9.1% for SFARI genes. On the other hand, the union of *InterVar* and *Psi-Variant* (*<u>I</u>* U *<u>P</u>*) achieved a diagnostic yield of 30.9% (Fig. 2C) (three times more than *<u>I</u>* ∩ *<u>P</u>*) but had a reduced effectiveness in detecting variants in SFARI genes (PPV = 0.141; OR = 3.18, 95% C.I. = 2.43–4.10) (Fig. 2A - 2B).

**Fig. 2.**
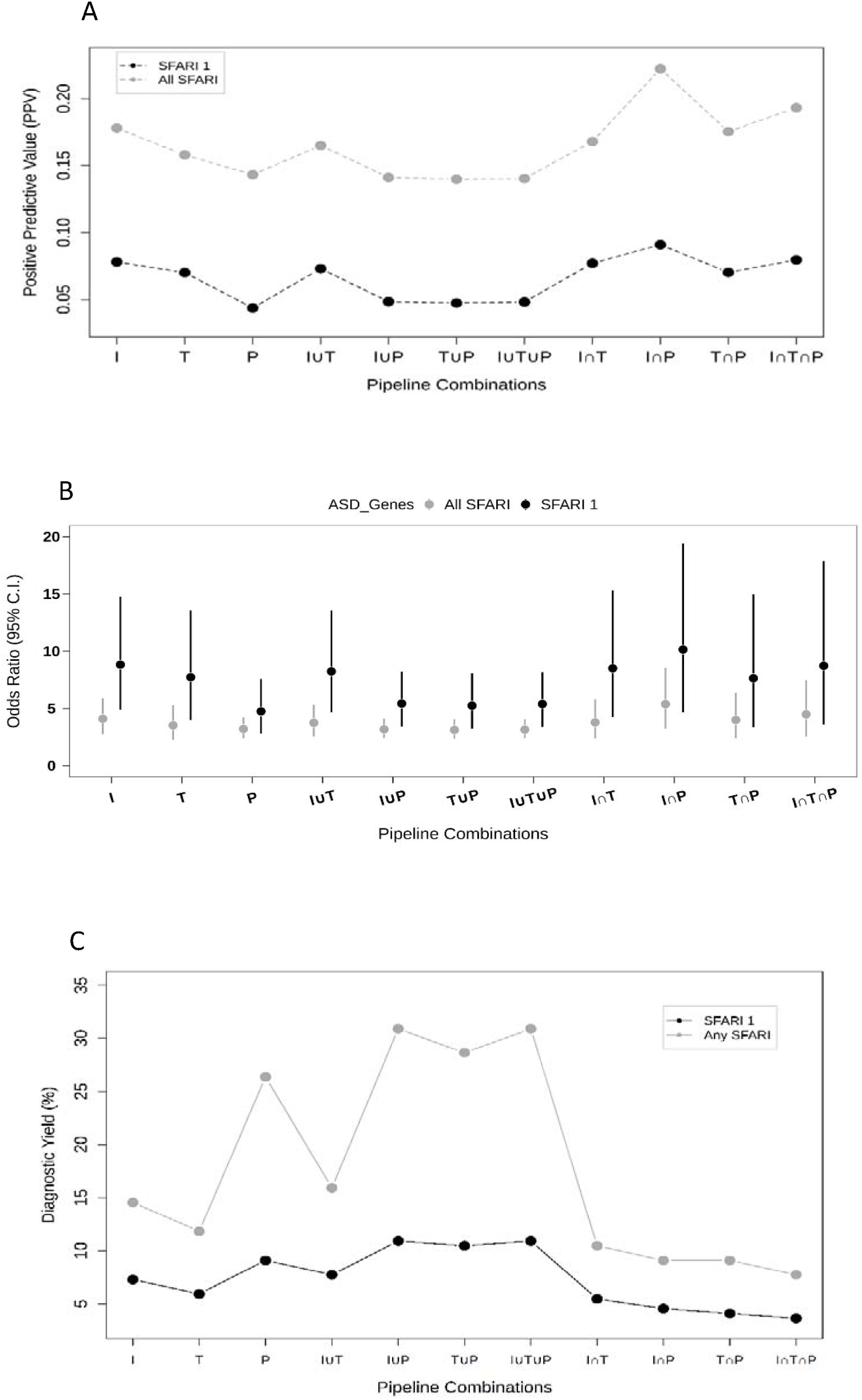
Effectiveness of *InterVar (I)*, *TAPES (T)*, *Psi-Variant (P)*, and their combinations in detecting candidate variants in ASD genes. **A** Positive predictive value (PPV) of detecting candidate variants in SFARI 1 and all SFARI genes. **B** Odds Ratios (ORs) of detecting candidate variants in SFARI 1 and all SFARI genes. **C** Diagnostic yield (%) achieved by the different tool combinations for detecting candidate variants in SFARI 1 and all SFARI genes.

## Discussion

In this study, we assessed the concordance and effectiveness of three bioinformatics tools in the interpretation of variants detected in the WES of children with ASD. There was better agreement in variant detection between *InterVar* and *TAPES* than between *Psi-Variant* and each of these two tools, probably because both *InterVar* and *TAPES* are based on the ACMG/AMP guidelines^8^, while *Psi-Variant* uses the interpretation of seven in-silico tools in assessing the functional consequences of LGD variants. In addition, most (94%) of the variants detected by either *InterVar* or *TAPES* were de-novo variants, compared to only 36% of the variants detected by *Psi-Variant*. This difference may be attributed to the fact that ACMG/AMP guidelines are particularly designed to detect de-novo highly penetrant variants, while inherited variants (autosomal recessive and X-linked) are usually classified as VUS^20^. Importantly, such rare inherited variants have been found to be associated with a variety of neurodevelopmental conditions, including ASD^9,17,18,21,22^. Another major difference between these tools lies in the detection of in-frame insertions/deletions that comprised ∼20% of the variants detected by either *InterVar* or *TAPES*, while such SNVs were discarded by *Psi-Variant*. We decided to exclude these variants from *Psi-Variant* because their clinical relevance has been demonstrated in several genetic disorders^37,38^ but not in ASD^39–41^.

Another important factor that could affect the concordance between the three tools is the annotation tools they use. Specifically, both *InterVar* and *TAPES* use AnnoVar^42^ for their variant annotation, while *Psi-Variant* uses Ensembl’s VEP^26^. It has already been shown that AnnoVar and VEP have a low concordance in the classification of LoF variants^43^. In addition, each tool, *InterVar*, *TAPES*, and *Psi-Variant*, utilizes a different set of in-silico tools for the classification of missense variants, with SIFT^29^ alone being shared by all three tools. These differences are probably the reason for the large differences in the detection of missense variants between the three tools (Table 1).

Today, there are no accepted guidelines for the detection of ASD susceptibility variants from WES data. Many genetic labs use the ACMG/AMP guidelines^8^, leading to a relatively low diagnostic yield^44,45^. Our findings suggest that different combinations of bioinformatics tools for variant interpretation may improve the detection of ASD susceptibility variants. Furthermore, combining these tools provides more flexibility in selecting the desired proportion between the detection yield and false positives. Thus, future guidelines for the detection of ASD susceptibility variants should consider the integration of different variant interpretation criteria.

Of note, many of the variants detected by the integrative pipeline affect genes with no known association with ASD, according to the SFARI Gene database^36^. This finding highlights the capability of the integrative pipeline to detect novel ASD genes. Obviously, the association of these genes and variants with ASD susceptibility needs to be validated in additional studies.

The results of this study should be considered under the following limitations. First, the effectiveness assessments of the different tools and their combinations were based on ASD genes from the SFARI Gene database^36^. While this is the most commonly used database for ASD genes and is continuously updated, it is based on data curated from the literature and may thus include genes falsely associated with ASD. Second, the variant detection analyses were performed on WES data of a cohort from the Israeli population, which may not necessarily be representative of the genetic architecture of ASD. Third, the tools used in this study were designed to detect only extremely rare variants with relatively large functional effects. Thus, a more effective approach for the detection of ASD susceptibility variants should also include the interpretation of other types of genomic variations, such as copy-number and compound heterozygote variants^46–51^, as well as other variants with milder functional effects^17,52,53^. Finally, it should be noted that there are many other approaches for variant interpretation from WES data. Thus, it is possible that combinations of other approaches would be more effective in the detection of ASD susceptibility variants from WES data than the approaches investigated in this study.

## Conclusions

Our findings suggest that combination of different bioinformatics tools is more effective in the detection of ASD candidate variants from WES data than each of the examined tools alone. Future guidelines for the detection of ASD susceptibility variants should consider integrating different variant interpretation approaches to improve the effectiveness of ASD candidate variants detection from whole exome sequencing data.

## Supporting information

Description of the machine learning false positive filter

List of variants detected by at least one of InterVar, Tapes, and Psi-Variant

## List of abbreviations

ACMG/AMP: American College of Medical Genetics and Genomics/Association of Molecular Pathology
ASD: autism spectrum disorder
C.I.: confidence interval
GATK: Genome Analysis Toolkit
LGD: likely gene disrupting
LoF: loss of function
LP: likely pathogenic
ML: machine learning
NADI: National Autism Database in Israel
NGS: next-generation sequencing
OR: odds ratio
P: pathogenic
PPV: positive predictive value
SNV: single nucleotide variants
VEP: Variant Effect Predictor
vcf: variant calling format
VUS: variants of uncertain significance
WES: whole exome sequencing.

## Declarations

### Institutional Review Board Statement

The study was conducted according to the guidelines of the Declaration of Helsinki and approved by the Ethics Committee of Soroka University Medical Center (SOR-076-15; 17 April 2016).

### Ethics approval and consent to participate

Informed consent was obtained from all the families involved in the study.

### Consent to publication

Not applicable.

### Availability of data and materials

WES data were generated as part of the ASC and are available in dbGaP with study accession: phs000298.v4.p3. The generated results and codes are available in a GitHub public repository: https://github.com/AppWick-hub/Psi-Variant or available upon reasonable request to the corresponding author Prof. Idan Menashe.

### Competing interests

The authors declare no competing interests.

### Funding

This study was supported by a grant from the Israel Science Foundation (1092/21).

### Authors’ contributions

*Conceptualization*: A.S. and I.M.; *methodology*: A.S. and I.M.; *software*: A.S. and L.L.; *validation*: A.S. and I.M.; *formal analysis*: A.S.; *resources*: H.G., G.M., A.M., Y.T., A.A. and I.D.; *data curation*: A.S.; *writing—original draft preparation*: A.S. and I.M.; *writing— review and editing*: I.M., and A.S.; *supervision*: I.M.; *project administration*: I.M.; *funding acquisition*: I.M. All the authors have read and agreed to the published version of the manuscript.

## Acknowledgments

We thank the families who participated in this research, without their contributions, genetic studies would be impossible.

## Notes

### Competing Interest Statement

The authors have declared no competing interest.

### Summary of Updates

Additional results of tool-specific diagnostic yield (highlighted in Fig. 2(C)) have been added. Also, a list of (Supplementary Table S2) LP/P/LGD variants (N=605) detected by at least one of InterVar, TAPES, and Psi-Variant, has been provided.

