## Supplementary material for "Comparison of three bioinformatics tools in the detection of ASD candidate variants from whole exome sequencing data": Description of the machine learning false positive filter

**Development of a Machine Learning (ML) false positive (FP) filter**

We developed an ML-based false positive (FP) classifier to filter out the FP SNVs in the downstream analysis. First, we prepared a truth set that consists of an outcome variable with two labels 0: Real SNVs; 1: Not real SNVs, and an array of variant quality-related metrics (DP, QD, QUAL, SOR, MQ, FS, etc.) for classifying a variant to be real or not. Each variant’s validation status (Real vs Not Real) was assigned after manually assessing these SNVs in the IGV.

In the truth set (Sample size=118), around 65% of the SNVs were not real. We checked the multi-collinearity issue and bivariate associations. Then the truth set was partitioned randomly based on our outcome variable into training and test sets with a probability of 0.65 for the training set (n = 78) and 0.35 for the testing set (n = 40). A five-fold cross-validation method was adopted to train the model.

Here two models, e.g., the multivariable logistic regression model and the Stochastic gradient boosting model, was trained. Diagnostic measures such as Area Under Curve (AUC) value, sensitivity, specificity, and misclassification error were computed and compared (highlighted in Table S1). We selected the Stochastic gradient boosting model over the logistic regression model based on the diagnostic measures. Further, we validated the best-fitted ML classifier on our testing set.

| **Table S1:** Model diagnostics on the training and testing set | | |
| --- | --- | --- |
| **Metrics** | **Training set** | **Testing set** |
| **Logistic regression model** | | |
| AUC | 0.9375 | 0.8874 |
| Sensitivity | 0.9253 | 0.8571 |
| Specificity | 0.9258 | 0.9615 |
| Misclassification Rate |  | 7.5% |
| **Stochastic Gradient Boosting Model** | | |
| AUC | 0.9567 | 0.9396 |
| Sensitivity | 0.9093 | 0.8571 |
| Specificity | 0.9298 | 1.0000 |
| Misclassification Rate |  | 5% |
